# Documentation and prevalence of prenatal and neonatal outcomes in a cohort of individuals with KBG syndrome

**DOI:** 10.1101/2023.01.17.23284321

**Authors:** Ola Kierzkowska, Kathleen Sarino, Drake Carter, Elaine Marchi, Anastassia Voronova, Gholson J. Lyon

## Abstract

Mutations in the Ankyrin Repeat Domain 11 (*ANKRD11*) gene are associated with KBG syndrome, a developmental disability that affects multiple organ systems and presents with a variety of skeletal, cardiac, gastrointestinal, and neuropsychiatric manifestations. The function of *ANKRD11* in human growth and development is not well understood, but its importance is established by the fact that constitutive or conditional gene knockout or mutation are lethal in mice embryos and/or pups. In addition, it plays a vital role in chromatin regulation, transcription, and development. Individuals with KBG syndrome are often misdiagnosed or undiagnosed until later in life, due to a combination of varying phenotypes, lack of targeted prenatal screening, and lack of documentation of prenatal and neonatal symptoms. The present study aims to report perinatal outcomes in individuals with KBG syndrome and their families and compare them to the prevalence in the overall population. We obtained data from 42 individuals through videoconferences, notes, and emails. Our results show that 45.2% of our cohort was born by C-section, 33.3% had a congenital heart defect, 23.8% were born before 37-weeks ‘ gestation, 23.8% were admitted to the NICU, 14.3% were small for gestational age, and 14.3% of the families in our cohort had a history of miscarriage. The prevalence of birth by C-section, premature birth, NICU admission, and small for gestational age was higher in our cohort compared to the overall population, including non-Hispanic and Hispanic populations. Other reports included feeding difficulties (21.4%), neonatal jaundice (14.3%), decreased fetal movement (7.14%), and pleural effusions *in utero* (4.67%). These findings suggest that there may be an association between *ANKRD11* variants or microdeletions and perinatal complications, and further research is needed to better assess the mechanisms behind this relationship. Comprehensive perinatal studies about KBG syndrome and updated documentation of its phenotypes can facilitate earlier detection, diagnosis, and treatment.

## Introduction

KBG syndrome is a rare genetic disorder that is caused by various mutations in the Ankyrin Repeat Domain 11 (*ANKRD11*) gene or microdeletions of chromosome 16q24.3. KBG syndrome affects multiple organ systems and can present with a wide range of phenotypes, including craniofacial and skeletal abnormalities, short stature, cardiovascular deformities, gastrointestinal issues, cognitive disabilities, developmental delay, seizures, and other clinical associations. Individuals with KBG syndrome are heterozygous for *ANKRD11* gene mutations,^1^ and mice with similar mutations also exhibit features of craniofacial abnormalities, reduced body size, and reduced bone mineral density.^2^

The effect of homozygous mutations in the *ANKRD11* gene in humans is unclear, although past studies indicate that homozygous mutations are lethal in mice embryos.^2^ Moreover, constitutive *ANKRD11* gene knockout has abnormal embryo development and pre-weaning lethality (www.mousephenotype.org).^3^ Notably, conditional knockout of *ANKRD11* in the neural crest, a transient embryonic tissue that contributes to the development of the skull, heart, and peripheral nervous system, results in neonatal lethality at least in part due to cleft palate and severely perturbed heart function^4^. Thus, while it is not currently clear when the constitutive *ANKRD11* knockout affects murine viability, it is likely that it causes embryo or early pup lethality. The lethality of mice that express mutated *ANKRD11* or mice with constitutive or conditional *ANKRD11* gene knockout demonstrates the importance of this gene in development even prior to birth, although the exact function of *ANKRD11* in human embryogenesis, growth, and development is yet to be uncovered. Two studies found that *ANKRD11* regulates neural stem cell proliferation and/or pyramidal neuron formation,^5,6^ while other studies illustrated the importance of *ANKRD11* in regulation of gene transcription and skeletogenesis.^2,4,6^ Furthermore, the exact causes and mechanisms responsible for producing mutations in *ANKRD11* are unknown. What is known is that at least some mutations in *ANKRD11* lead to *ANKRD11* protein instability, as well as altered transcriptional activity and epigenetic signature.^6–9^ As with all diseases, the intersection of genetics, epigenetics, and environmental factors is critical in determining etiology.^10^ In the case of a rare disorder such as KBG syndrome, little is known about the combination of factors that produce it or whether any genetic and environmental insults can manifest as early as during pregnancy.

The present study aims to document prenatal, birth, and postnatal outcomes and complications in individuals with KBG syndrome to further uncover the relationship between *ANKRD11* mutations and human development, as well as provide updated data for future prenatal or neonatal screening. To explore the importance of our findings, we compared the prevalence of perinatal outcomes in our cohort to the prevalence of these outcomes in the general population. There is only one report of a prenatal diagnosis of KBG syndrome,^11^ and our work seeks to cumulatively report perinatal manifestations in individuals with KBG syndrome.

## Methods

The New York State Psychiatric Institute – Columbia University Department of Psychiatry Institutional Review Board approved this study. 42 individuals (18 females, 24 males) from 37 families were interviewed via Zoom by a single clinician (G.J.L) from February 2021 to May 2022. Data collected from 25 of these individuals has already been published.^12^ The individuals interviewed reside in 11 different countries and were recruited via a private Facebook group created by the KBG Foundation or by self-referral. Inclusion criteria included a molecular diagnosis of KBG syndrome, which was confirmed by review of medical records by G.J.L. Informed consent to disclose health information and photographs and to record each interview was obtained prior to videoconferencing. Genetic reports, medical records, and photographs were collected from the families through email and compiled before the interviews. The duration of the interviews were approximately 1-2 hours long and consisted of the physician (G.J.L) asking structured questions and visually assessing participants for phenotypic facial and limb characteristics. Interview questions aimed to gain a comprehensive medical, surgical, familial, prenatal, and developmental history. The physician recorded written notes throughout the videoconference which were then summarized into summary tables.

To compile data specifically about pregnancy complications and the participants ‘ prenatal and postnatal trajectories, information was taken from the summary tables of each videoconference with a focus on number of miscarriages, gestational age of the participant at birth, and the presence or absence of prenatal, birth, and postnatal complications. Data was entered into Microsoft Excel to organize and further analyze the findings. Some videoconferences did not specifically include in-depth information about the participant ‘s prenatal period; therefore, emails were sent to all 37 families requesting further information. The families were reassured that all their responses were voluntary and confidential. Questions in the email included 1) History and number of miscarriages, 2) Use of fertility treatments and why they were needed, 3) Complications during pregnancy that suggested the participant may have a genetic syndrome, 4) Any other relevant information about the pregnancy, birth, or postnatal period that the parent wanted to share. The information gathered from the emails was also entered into Microsoft Excel along with the initial summary data. 12 out of 37 families (32.4%) responded to the email with additional details about the pregnancy with and birth of their child(ren) with KBG syndrome. The presence of a trait or phenotype was documented as such when it was explicitly stated in the interview, found in the individual ‘s medical records, or stated in email. The absence of a trait was documented as such if the interviewee stated that they did not possess it, if it was not mentioned within the interview, and/or if it was not found in the medical records.

The perinatal events of interest included history of miscarriage, birth by cesarian section (C-section), premature birth (<37 week gestation), small for gestational age, decreased fetal movement *in utero*, NICU admission after birth, congenital heart defects, neonatal jaundice, feeding difficulties, failure to thrive, respiratory complications, and nuchal translucency. The number of individuals that experienced each of these complications were added together for a total number of people per complication. All percentages were calculated out of 42 (total number of individuals within our cohort). Because the majority of our cohort identified as either Caucasian non-Hispanic or Caucasian Hispanic, we further analyzed our data based on these two ethnicities. This was done to assess if there were similarities or differences in perinatal outcomes between ethnicities, as past evidence suggests individuals of Hispanic, Black, and American Indian ethnicity and racial background have increased rates of adverse pregnancy outcomes.^13^ Our non-Hispanic Caucasian participants included those that reside in Spain and Portugal.

To compare the prevalence of pregnancy outcomes in our cohort to those of the overall population, a literature review was performed. PubMed and Google Scholar were searched for studies published between 2000 and 2022. Studies were included if they provided the epidemiology, rate, or prevalence of each perinatal event (I.e: premature birth, crude NICU admission rates, neonatal jaundice, etc.) globally or within the United States or Europe, as this is where the majority of our cohort resides (**Table 2**). Studies were excluded if the prevalence of each perinatal event was not stated, if the study focused on a region of the world outside of the United States or Europe, or if the sample size was less than 5,000 participants (**Figure 1**). Between 2 and 5 papers per pregnancy outcome were found, giving a total of 12 papers used (**Table 1**).^14–25^ Once all of the papers for each outcome were selected, the average prevalence was calculated to have one number to compare our prevalence to. The papers with their sample sizes, year of study, region of study, and summarized findings are listed in **Table 1**.

**Figure 1:**
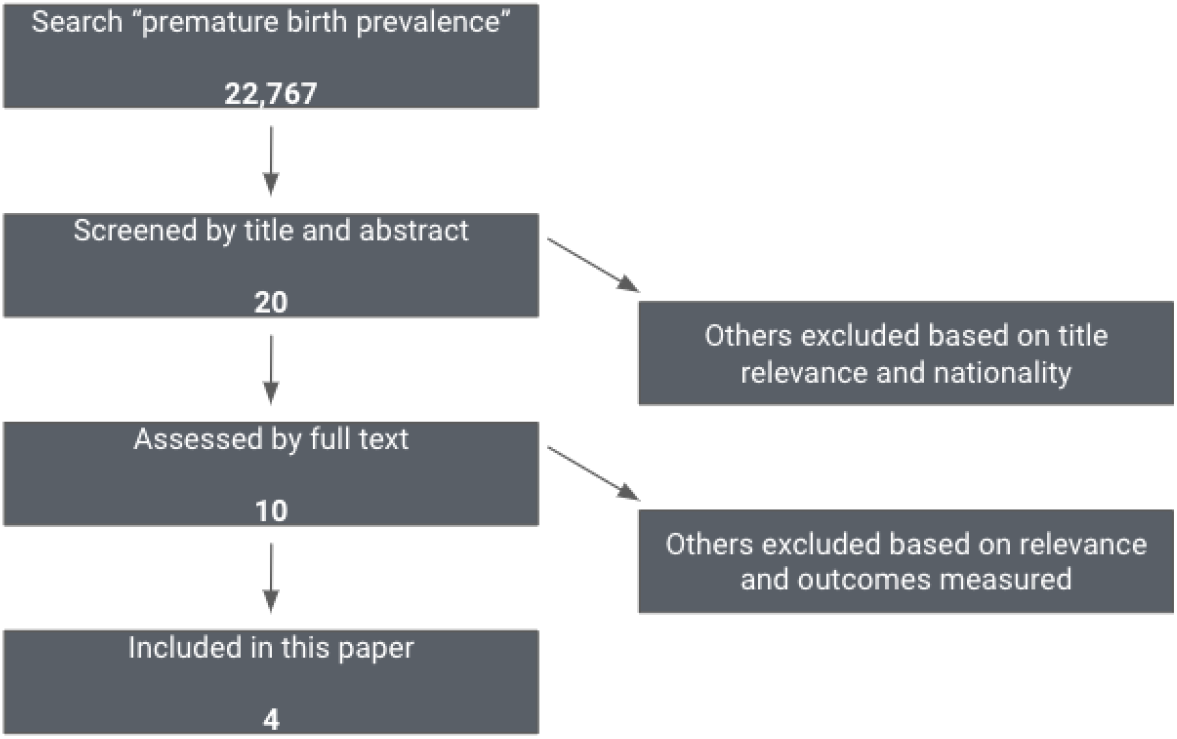
Example flowchart for inclusion and exclusion of comparison papers Demographics.

**Table 1:**
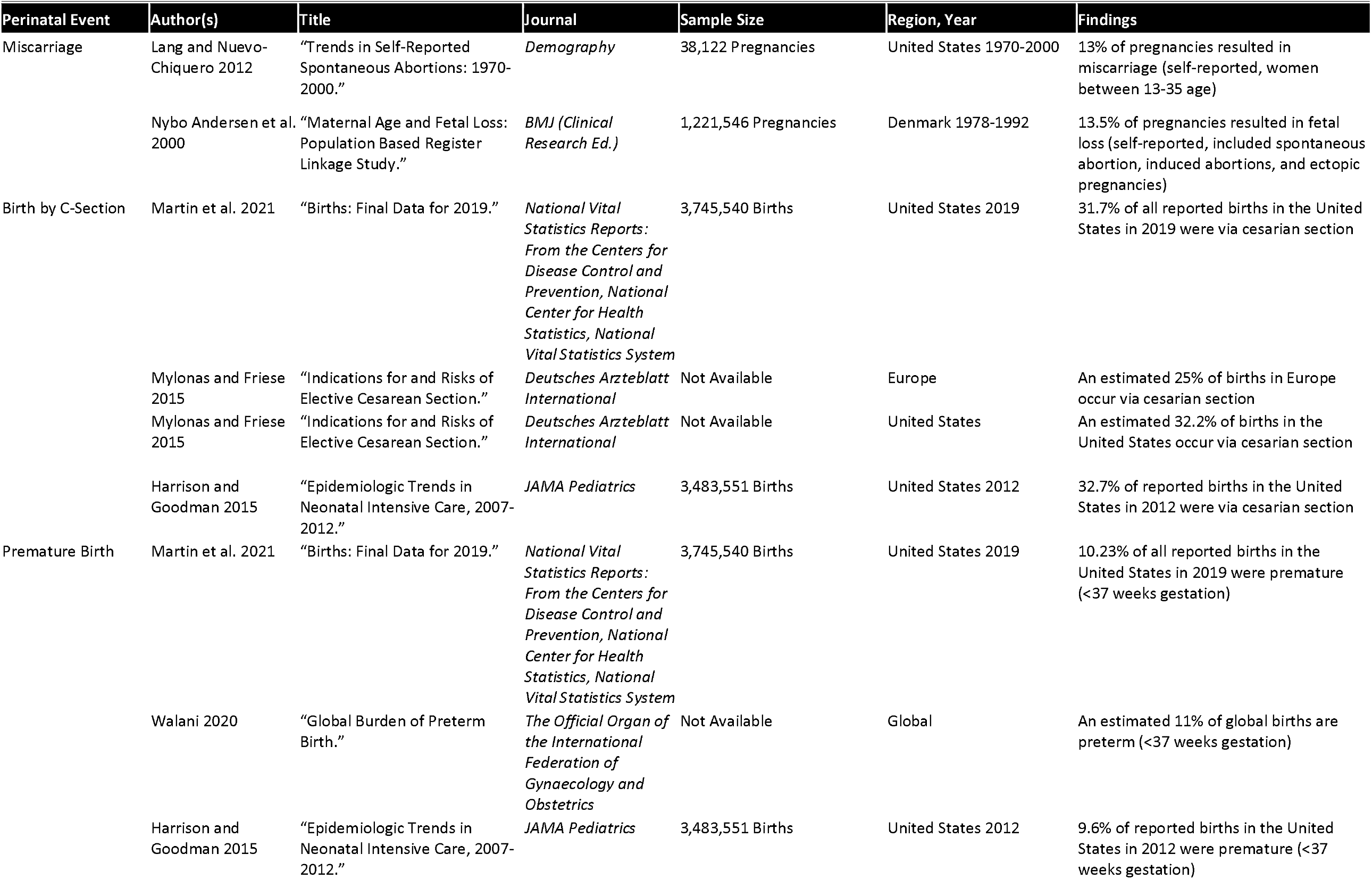
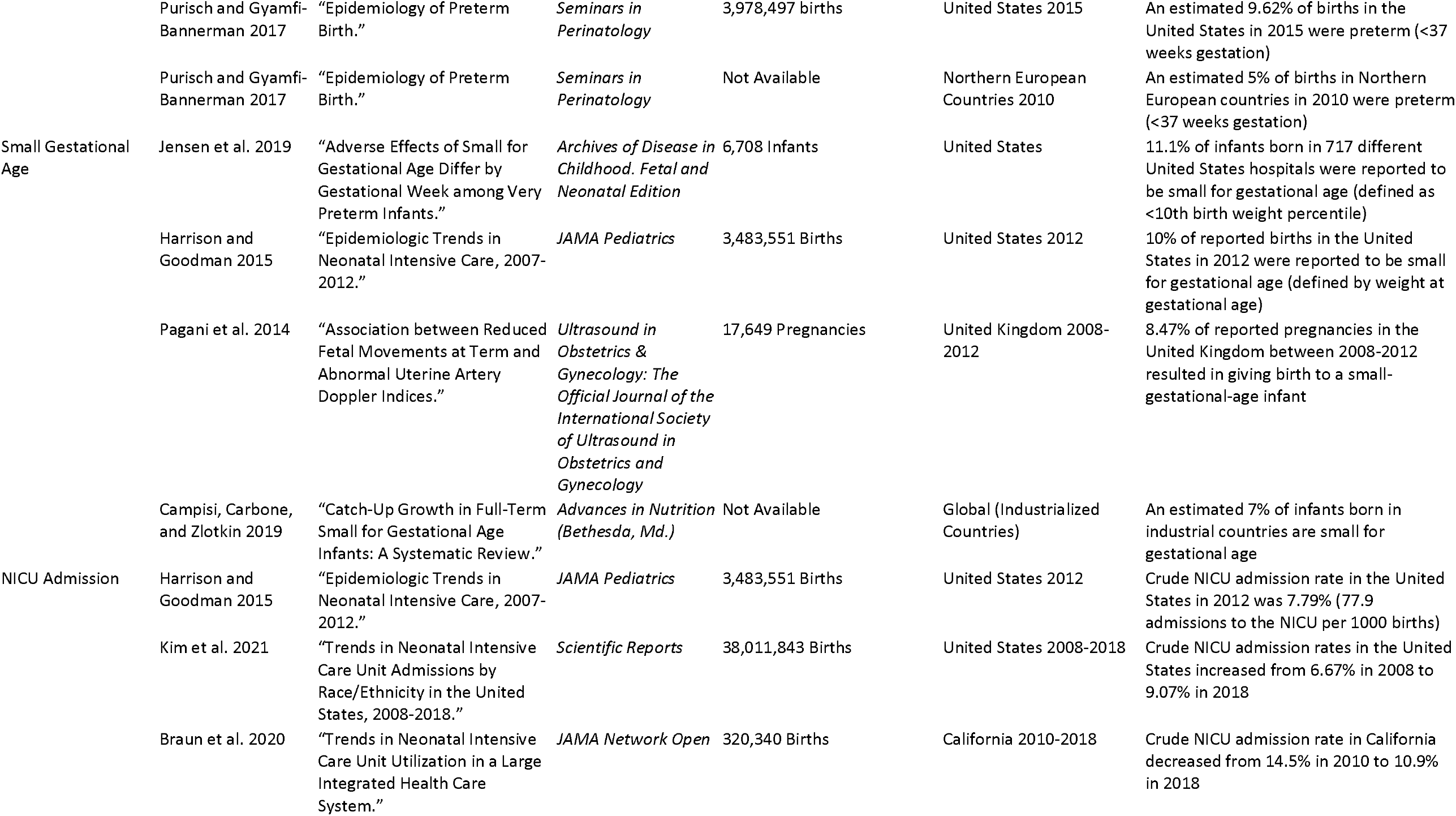
Summary table of comparison papers with sample sizes and findings

## Results

Demographic information for our cohort is summarized in **Table 2**. Out of 42 individuals, the average age was 13.64 (range: 1-61). 98% of our cohort identified as Caucasian, with 85.7% identifying as non-Hispanic and 14.3% identifying as Hispanic. 57.1% of our cohort lives in the United States and 28.6% of our cohort lives in a European country. **Figure 2** demonstrates the prevalence of all perinatal events of interest within our cohort. These pregnancy outcomes are discussed below. **Table 3** presents the comparisons between our cohort and the overall population. History of miscarriage, birth by C-Section, premature birth, NICU admission, and small gestational age were chosen because these 5 variables had the greatest amount of information in the literature to make the best comparisons to. Our cohort had increased total average rates of miscarriage (14.3% vs 13.1%), birth by C-Section (45.2% vs 30.4%), prematurity (23.8% vs 9.09%), NICU admission after birth (23.8% vs 9.25%), and small for gestational age (14.3% vs 9%) compared to the overall population. **Table 4** displays the comparisons between our cohort ‘s non-Hispanic population and overall non-Hispanic populations. Within our cohort of 36 non-Hispanic individuals, there were increased rates of all 5 outcomes compared to the overall populations. **Table 5** shows the comparisons between our cohort ‘s Hispanic population and overall Hispanic population. Within our cohort of 6 Hispanic individuals, there were increased rates of C-section, prematurity, and NICU admission after birth compared to the overall Hispanic population. Our Hispanic population did not have a history of miscarriages, although the rate in overall Hispanic populations was found to be 1.56%. Data could not be found for rates of small gestational age in overall Hispanic populations.

**Table 2:** Cohort Demographics

PERINATAL OUTCOMES IN KBG SYNDROME
| Table 2: Cohort Demographics |  |  |
| --- | --- | --- |
| Factor | Level | Number |
| Total |  | 42 |
| Sex | Male | 24 (57.1%) |
|  | Female | 18 (42.9%) |
| Average Age (range) |  | 13.64 (1, 61) |
| Race | Caucasian | 41 (98%) |
|  | Asian | 1 (2%) |
| Ethnicity | Non-Hispanic | 36 (85.7%) |
|  | Hispanic | 6 (14.3%) |
| Country of Origin | United States | 24 (57.1%) |
|  | Spain | 5 (11.9%) |
|  | United Kingdom | 4 (9.52%) |
|  | Germany | 2 (4.76%) |
|  | Canada | 1 (2.38%) |
|  | Mexico | 1 (2.38%) |
|  | Australia | 1 (2.38%) |
|  | Portugal | 1 (2.38%) |
|  | Argentina | 1 (2.38%) |
|  | Ecuador | 1 (2.38%) |
|  | Lebanon | 1 (2.38%) |

**Figure 2:**
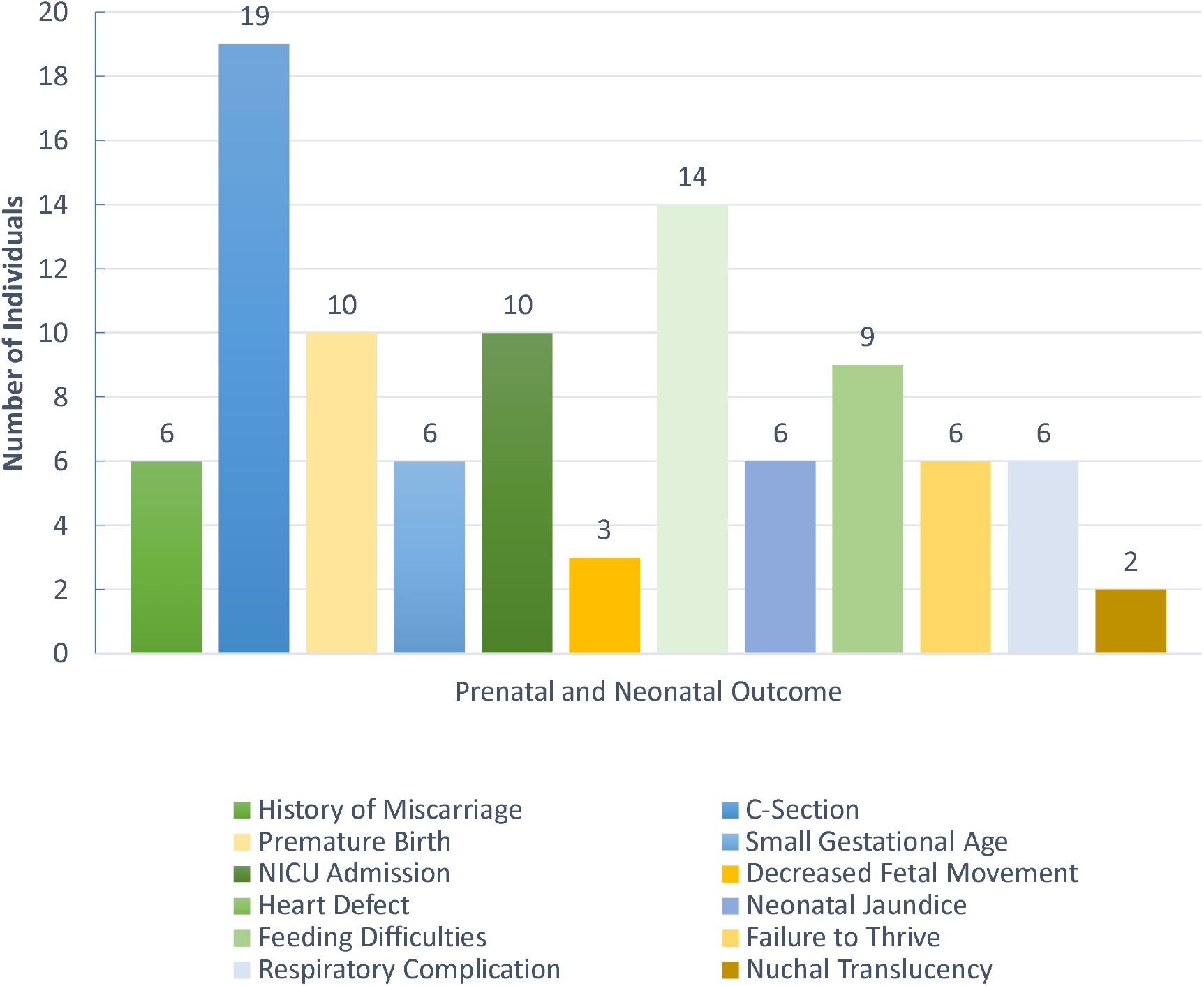
Prevalence of Perinatal Events in our cohort. Each colored bar indicates the number of individuals in our cohort (out of the total sample size, 42) with a specific prenatal and neonatal outcome.

**Table 3:**
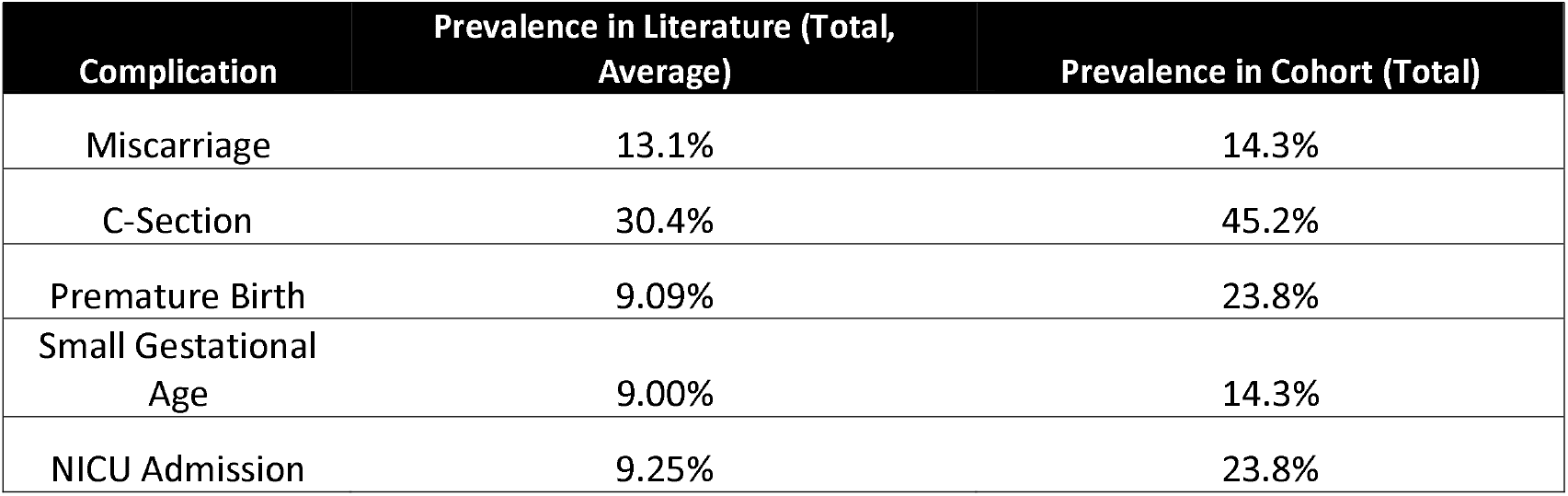
Comparing overall prevalence to KBG syndrome cohort

| Complication | Prevalence in Literature (Total, Average) | Prevalence in Cohort (Total) |
| --- | --- | --- |
| Miscarriage | 13.1% | 14.3% |
| C-Section | 30.4% | 45.2% |
| Premature Birth | 9.09% | 23.8% |
| Small Gestational Age | 9.00% | 14.3% |
| NICU Admission | 9.25% | 23.8% |

**Table 4:** Comparing non-Hispanic population prevalence to KBG syndrome cohort

| Complication | Prevalence in Literature (non-Hispanic Caucasian) | Prevalence in Cohort (non-Hispanic Caucasian) |
| --- | --- | --- |
| Miscarriage | 9.23% | 16.7% |
| C-Section | 30.7% | 41.7% |
| Premature Birth | 9.26% | 22.2% |
| Small Gestational Age | 8.50% | 13.9% |
| NICU Admission | 8.50% | 25.0% |

**Table 5:**
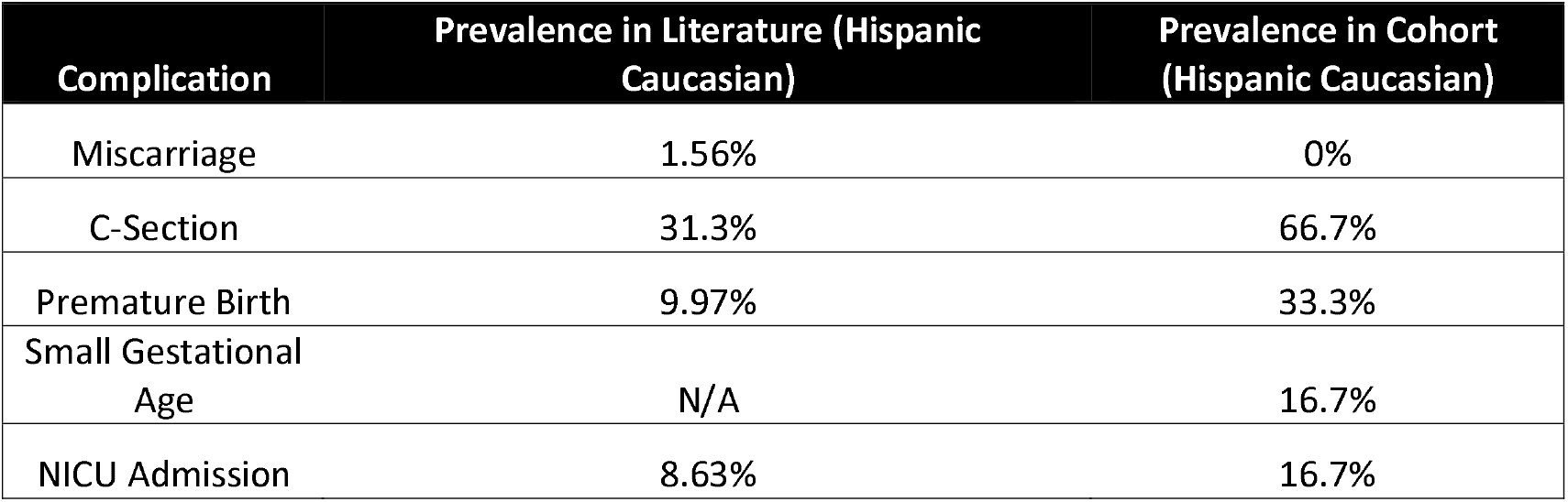
Comparing Hispanic population prevalence to KBG syndrome cohort

| Complication | Prevalence in Literature (Hispanic Caucasian) | Prevalence in Cohort (Hispanic Caucasian) |
| --- | --- | --- |
| Miscarriage | 1.56% | 0% |
| C-Section | 31.3% | 66.7% |
| Premature Birth | 9.97% | 33.3% |
| Small Gestational Age | N/A | 16.7% |
| NICU Admission | 8.63% | 16.7% |

Of the families that reported having a C-section, 57.9% of them were due to an emergency situation. 26.3% of C-sections were due to fetal distress, 21.1% of C-sections were due to breech position or being stuck in the birth canal, and 10.5% of C-sections were due to heart decelerations or undetectable heart rate during delivery.

Our cohort reported other important findings that did not have a corresponding prevalence in the literature. When our cohort was asked if there were any pregnancy complications that suggested to their doctors or clinicians that their child may have a genetic syndrome, the complications that were reported most frequently were small gestational age (14.3%) and decreased fetal movement (7.14%) (**Figure 2**). Two families (4.67%) reported their children having persistent left superior vena cava syndrome. Two families (4.67%) reported their children having a type of pleural effusion *in utero*. One of these patients was diagnosed with mild pleural effusion at 28 weeks gestation, while the other patient was diagnosed with bilateral chylothorax at 28 weeks gestation. This compromised fetal wellbeing and the latter patient underwent *in utero* shunt placement surgery at 33 weeks ‘ gestation. The newborn continued to have pleural effusions until 6 weeks of age. In addition, one family reported their child having bleeding choroid plexus cysts in the brain that resolved. In this case, the doctors suspected the child had Edwards syndrome, but this was subsequently ruled out. Two families (4.67%) reported their children having increased nuchal translucency during pregnancy (**Figure 2**), although the definition of nuchal translucency by their doctors could not be confirmed.

Due to the heterogeneity of KBG syndrome presentations, other findings were reported including 33.3% of participants with a congenital heart defect that presented *in utero* or after birth (**Figure 2**). These heart defects included five participants with an atrial septal defect or ventricular septal defect, the two participants with left superior vena cava syndrome, three participants with a patent foramen ovale, one participant with a patent ductus arteriosus, and one participant with tetralogy of Fallot. 14.3% of participants had a respiratory defect upon birth (**Figure 2**), including cyanosis, apnea, hypoxia, and neonatal respiratory distress syndrome. Of these, one participant presented to the NICU with cyanosis on delivery and hypoxia along with low temperature and neonatal jaundice. Prader Willi syndrome was suspected at this time but was subsequently ruled out. Another participant was initially diagnosed with Cornelia de Lange syndrome but this too was subsequently ruled out.

14.3% of our participants experienced neonatal jaundice. This is compared to the 50-80% reported in the literature for overall populations, including term and preterm infants, respectively.^26^ 21.4% of our participants experienced feeding difficulties in infancy and young childhood. This can be compared to 42% for premature infants, 20-50% for term children, and 70-89% for individuals with developmental disabilities.^27,28^ 14.3% of our participants experienced failure to thrive (**Figure 2**).

There are a few factors to consider regarding our participants ‘ mothers and their experiences during pregnancy. One family reported taking several medications during pregnancy, including corticosteroids (Prednisone and Betamethasone), Valium for contractions, Ventolin to stop early contractions, Penicillin for a Group B Strep infection, and Adalat to slow down her heart rate. Of note, this family was expecting quadruplets. Only one of the four children has KBG syndrome. In addition, one family reported maternal epilepsy treated with Topiramate during pregnancy, one family reported maternal diabetes also treated during pregnancy, and one family reported being prescribed magnesium sulfate at 29 weeks ‘ gestation due to preterm labor.

## Discussion

The most common perinatal complication reported by our cohort was birth by C-section. Although this is not a complication or adverse event in itself, the majority of C-sections in our cohort were due to birth complications. The second most common complications reported by our cohort were heart defects, premature birth (<37 weeks), and NICU admission immediately after birth. Other common complications included feeding difficulties in infancy, small gestational age *in utero*, a history of miscarriage, neonatal jaundice, failure to thrive in infancy, and respiratory complications shortly after birth. In our cohort, KBG syndrome manifested itself in a variety of different ways, including pleural effusions, choroid cysts, and various heart defects such as superior vena cava syndrome and patent foramen ovale.

The prevalence of miscarriage, premature birth, C-sections, small gestational age, and NICU admission were higher in our cohort compared to the overall population. The prevalence of neonatal jaundice and feeding difficulties in infancy were lower in our cohort compared to the overall population. More research is needed to confirm rates of these complications in the general population. The increased rates of perinatal complications in a cohort of individuals with KBG syndrome suggests that there may be an association between *ANKRD11* mutations and adverse outcomes in fetuses and newborns. The cardiac deficits and small gestational age findings are consistent with past reports;^11,29^ however, the other findings presented here have not been previously documented.

The limitations of this study are as follows: A sample size of 42 individuals from 37 families does not provide adequate power to confirm statistical significance of these findings. Likewise, a larger sample of Hispanic participants with KBG syndrome is needed to confirm whether rates of adverse perinatal events are higher than the overall Hispanic population. In addition, statistical significance between cohort data and population data could not be analyzed due to unknown statistical tests used by the other authors. An interesting limitation to note is that 12 families responded to email inquiries. These families may have responded with additional information because the inquiry applied to their situation. While this demonstrates a bias in responses, our data was gathered from videoconference notes; the email responses served as an adjunct. Other limitations involve the use of past studies: some of the epidemiological papers provided estimates for prevalence or defined variables differently. For example, the papers included in this study defined small gestational age at birth using birth weight while we defined small gestational age as a subjective report by a participant ‘s provider while the child was *in utero*. In addition, the majority of our results were reported subjectively by our participants, as some families did not provide medical records. It would be ideal to review objective data from medical records, if and when we can ever obtain such records, although this has proven to be difficult as many families do not have the records to provide. Finally, it is not clear whether there is a relationship between maternal medication use, such as those described, and perinatal outcomes.

Limitations aside, this study further documents prenatal and neonatal phenotypes of KBG syndrome. There is one documented case of prenatal confirmation of KBG syndrome, in which a chromosomal microassay (CMA) demonstrated a fetus with a deletion of chromosome 16, including *ANKRD11*^11^; however, more studies are needed to further elucidate the prenatal and neonatal phenotypes of KBG syndrome to create more targeted screening measures. More often than not, individuals with KBG syndrome are misdiagnosed or undiagnosed until later in life.^1^ This is in part due to KBG syndrome ‘s varying phenotypes, which are sometimes considered clinically mild or bear few complications in the neonatal period.^11^ It is also in part due to the lack of targeted prenatal testing and the lack of documentation of prenatal and neonatal signs and symptoms. Our report suggests the medical community needs to continue to document ultrasound findings, pregnancy complications, and postnatal complications. This, in conjunction with chromosomal microassay and exome sequencing, can facilitate prenatal and postnatal counseling, allow more accurate and earlier diagnosis, and support both families and medical personnel in delivering and caring for infants with KBG syndrome across the world.

## Data Availability

All data produced in the present study are available upon reasonable request to the authors.

## Acknowledgements

We would like to thank all of the KBG syndrome families who participated in this study, along with the KBG Foundation for referrals.

## Author Contributions

GJL was responsible for all videoconferencing and primary data collection, with secondary summaries performed by members of the team including EM. Data analysis was performed by OK. The first draft of the manuscript was written by OK, with extensive editing thereafter by GJL, KS, DC and AV.

## Ethical Approval

Both oral and written patient consent were obtained for research and publication, with approval of protocol #7659 for the Jervis Clinic by the New York State Psychiatric Institute - Columbia University Department of Psychiatry Institutional Review Board.

